# Viral Load of SARS-CoV-2 in Respiratory Aerosols Emitted by COVID-19 Patients while Breathing, Talking, and Singing

**DOI:** 10.1101/2021.07.15.21260561

**Authors:** Kristen K. Coleman, Douglas Jie Wen Tay, Kai Sen Tan, Sean Wei Xiang Ong, Than The Son, Ming Hui Koh, Yi Qing Chin, Haziq Nasir, Tze Minn Mak, Justin Jang Hann Chu, Donald K. Milton, Vincent T. K. Chow, Paul Anantharajah Tambyah, Mark Chen, Tham Kwok Wai

**Author notes:** **Corresponding Author:** Tham Kwok Wai, PhD; Address: 4 Architecture Drive, Singapore 117566. **Alternate Corresponding Author:** Kristen K. Coleman, PhD; Address: 8 College Rd, Level 9, Singapore 169857. Authors KKC, DJWT, KST, and SWXO contributed equally. Authors TTS, MHK, YQC, HN, and TMM contributed equally.

## Abstract

**Background:** Multiple SARS-CoV-2 superspreading events suggest that aerosols play an important role in driving the COVID-19 pandemic. However, the detailed roles of coarse (>5μm) and fine (≤5μm) respiratory aerosols produced when breathing, talking, and singing are not well-understood.

**Methods:** Using a G-II exhaled breath collector, we measured viral RNA in coarse and fine respiratory aerosols emitted by COVID-19 patients during 30 minutes of breathing, 15 minutes of talking, and 15 minutes of singing.

**Results:** Among the 22 study participants, 13 (59%) emitted detectable levels of SARS-CoV-2 RNA in respiratory aerosols, including 3 asymptomatic patients and 1 presymptomatic patient. Viral loads ranged from 63–5,821 N gene copies per expiratory activity per patient. Patients earlier in illness were more likely to emit detectable RNA, and loads differed significantly between breathing, talking, and singing. The largest proportion of SARS-CoV-2 RNA copies was emitted by singing (53%), followed by talking (41%) and breathing (6%). Overall, fine aerosols constituted 85% of the viral load detected in our study. Virus cultures were negative.

**Conclusions:** Fine aerosols produced by talking and singing contain more SARS-CoV-2 copies than coarse aerosols and may play a significant role in the transmission of SARS-CoV-2. Exposure to fine aerosols should be mitigated, especially in indoor environments where airborne transmission of SARS-CoV-2 is likely to occur. Isolating viable SARS-CoV-2 from respiratory aerosol samples remains challenging, and whether this can be more easily accomplished for emerging SARS-CoV-2 variants is an important enquiry for future studies.

**Key Points:** We sampled respiratory aerosols emitted by COVID-19 patients and discovered that fine aerosols (≤5μm) generated during talking and singing contain more SARS-CoV-2 copies than coarse aerosols (>5μm) and may play a significant role in the transmission of SARS-CoV-2.

## Introduction

Coronavirus disease 2019 (COVID-19) is caused by the highly transmissible severe acute respiratory syndrome coronavirus 2 (SARS-CoV-2). Irrespective of symptomatology, COVID-19 patients can harbor high viral loads of SARS-CoV-2 in their respiratory tracts [1, 2], and emit SARS-CoV-2 RNA into the air [3, 4], which may be culturable under favorable circumstances and collection methods [5]. Although virus emissions from talking and singing have not been measured, these expiratory activities are hypothesized to play a crucial role in virus transmission [6]. A significant proportion of SARS-CoV-2 transmission is estimated to be from asymptomatic individuals [7], and multiple SARS-CoV-2 superspreading events [8-10] suggest that aerosols may be critical in driving the COVID-19 pandemic. Thus, refined public health measures are likely needed to contain the virus, especially in under-vaccinated populations.

Respiratory aerosols range from 0.1–100μm in diameter and can be categorized as coarse (>5μm) and fine (≤5μm) aerosols, based on where they deposit in the respiratory tract [11]. Coarse aerosols are inhalable and deposit in the upper airways, whereas fine aerosols are respirable and deposit in the lower airways. The amount of infectious virus these size fractions carry and their relative importance to SARS-CoV-2 transmission and infection is not well-understood. Experimental studies of non-human primates have demonstrated that COVID-19 may be anisotropic [12] as more severe illness results from inhaling infectious aerosols that are 1–3μm in diameter when compared to direct intranasal and intratracheal inoculation [13]. Other models, however, demonstrate a disease spectrum similar to humans with combined intranasal and intratracheal inoculation [14]. Cynomolgus macaques also shed more SARS-CoV-2 in fine aerosols when compared to coarse aerosols [15]. To better understand the detailed role of respiratory aerosols in the transmission of SARS-CoV-2, we measured viral loads in coarse and fine respiratory aerosols emitted by COVID-19 patients during breathing, talking, and singing.

## Materials and Methods

### Patient Recruitment and Data Collection

Participants were recruited from February–April 2021 at the National Centre for Infectious Diseases in Singapore. All newly admitted patients were screened based on the following inclusion criteria: age ≥21 years and positive for COVID-19 via reverse transcription-quantitative polymerase chain reaction (RT-qPCR). Basic demographic data were recorded. Symptom data were collected based on a list of seven pre-specified symptoms. For asymptomatic individuals, the day of diagnosis was recorded as day one of illness. Cycle threshold (Ct) values of clinical respiratory samples and SARS-CoV-2 serology test results were obtained from medical records. Virus genome sequence data were obtained from National Public Health Laboratory records.

### Expiratory Sample Collection

Expiratory samples were collected using the G-II exhaled breath collector, described in detail by McDevitt *et al*. [16]. Briefly, study participants were seated facing the truncated cone-shaped inlet, with air drawn continuously around the subject’s head and into the sampler (Figure 1). The cone served as a capture type ventilation hood which allowed the collection of expiratory particles with minimal fugitive emissions. Participants were asked to perform three separate expiratory activities on the same day: 30 minutes of tidal breathing, 15 minutes of talking, and 15 minutes of singing. For the talking activity, participants were asked to repeat passages read to them from the children’s book, “Green Eggs and Ham” by Dr. Seuss. For the singing activity, participants were asked to sing “Happy Birthday”, “ABC song”, “Twinkle, Twinkle, Little Star”, and “We Wish You a Merry Christmas” with background music. Aerosols were collected in two size fractions, namely coarse (>5μm) and fine (≤5μm). The coarse fraction was collected on a Teflon® surface as intake air (130L/min) flowed through a conventional slit impactor. The Teflon® impactor was swabbed thrice, end to end, with a flocked swab first dipped in 1× phosphate buffered saline (PBS) solution with 0.1% bovine serum albumin (BSA). The swab was rotated during swabbing to ensure that all surfaces of the flocked tip were in contact with the impactor for optimal retrieval of coarse particles. The flocked swab was then placed in a 15mL conical tube containing 1mL of 1× PBS with 0.1% BSA. Fine particles were condensed into a reservoir of 1× PBS with 0.1% BSA and collected into 50mL conical tubes. In between each activity, the G-II was decontaminated with 10% bleach, rinsed with water, and wiped dry.

**Figure 1.**
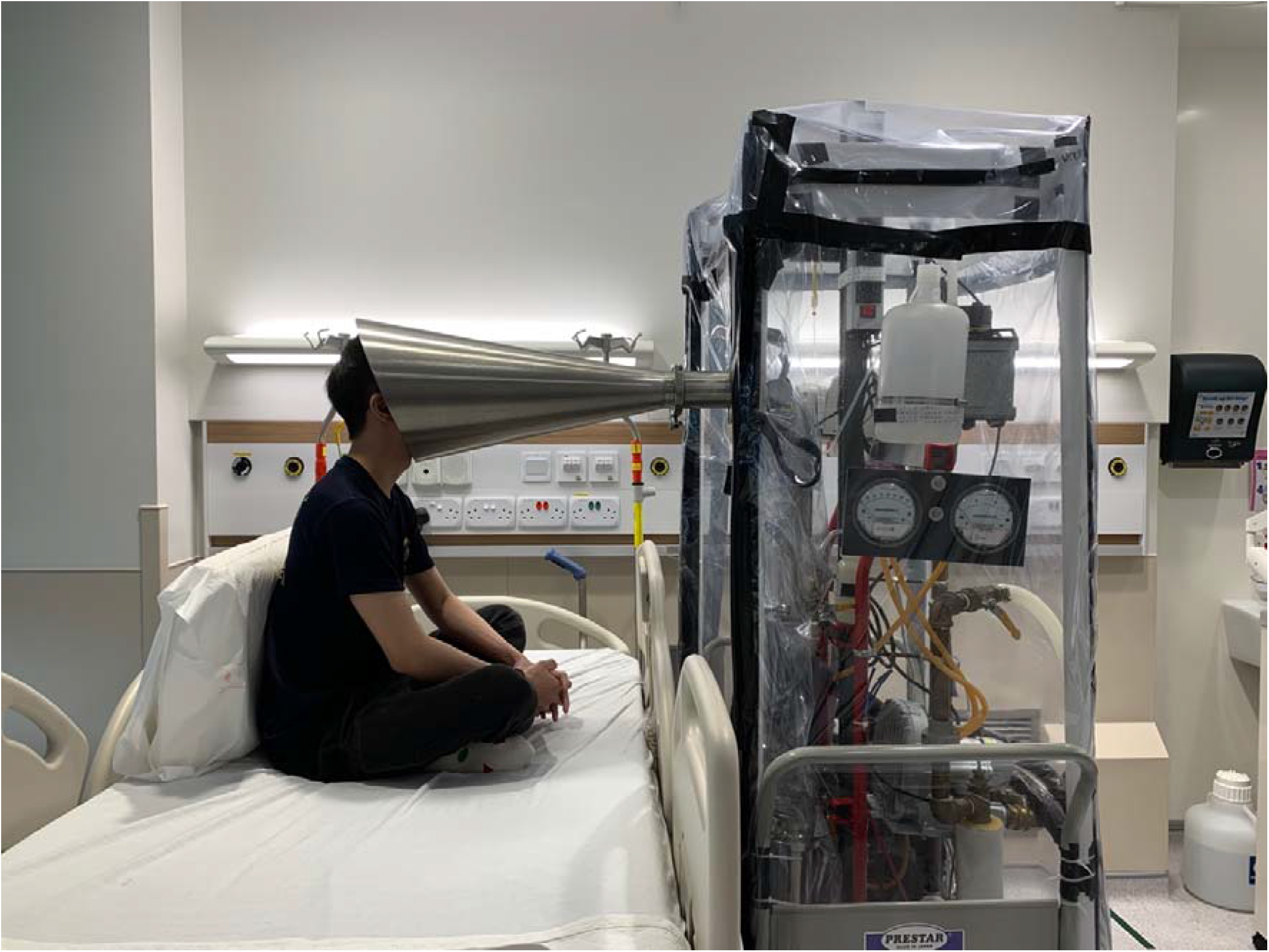
Schematic representation of expiratory sample collection using the G-II exhaled breath collector inside the COVID-19 patient room.

### Sample Processing

Samples were transported to and processed in the National University of Singapore Biosafety Level 3 Laboratory on the same day as collection. Coarse fraction samples were vortexed for 15 seconds, and aliquoted into 1.5mL screw-capped tubes. Fine fraction samples were concentrated with two to four Amicon Ultra-15 100 kDA centrifugal filter units (Millipore, USA), pooled, and filtered through a 0.22μm centrifuge tube filter (Corning). The filtrate was topped up to 1.6mL with Dulbecco’s Modified Eagle Medium (DMEM) supplemented with 2% fetal bovine serum (FBS) and 1× antibiotic-antimycotic. Fine fraction samples were used for viral culture on the same day, with the remaining volumes aliquoted into 1.5mL screw-capped tubes. Coarse fraction samples were not cultured as the impaction method was not designed for culture analysis [16]. Samples were kept at - 80°C prior to RNA extraction.

### Virus Culture

Vero E6 cells were cultured in DMEM supplemented with 2% FBS in a 5% CO_2_ humidified incubator at 37°C. Cells were seeded in T25 flasks at 1 to 2 days prior to sample inoculation. Flasks were replaced with fresh media (DMEM with 2% FBS and 1× antibiotic-antimycotic) and inoculated with 750µl of each processed sample (culture 1). Presence of cytopathic effect (CPE) was monitored every 3 to 5 days, and a subsequent passage was performed on day 7 (culture 2). Cultures 1 and 2 were observed for up to 14 days each, and supernatants were harvested at days 7 and 14 for RT-qPCR.

### Viral RNA Quantification

RNA was extracted from 200μl of each expiratory sample using the QIAamp MinElute Virus Spin Kit (Qiagen, Germany) according to the manufacturer’s instructions. RNA was eluted in 50μl of nuclease-free water (NFW) and stored at -80°C. The CDC N1 assay (Centers for Disease Control and Prevention, USA) was performed for the detection of SARS-CoV-2. A 20µl reaction mix was prepared with 5µl of TaqPath™ 1-Step RT-qPCR Master Mix, CG (Thermo Fisher Scientific, USA), 1.5µl of primer-probe mix, 3.5µl of NFW, and 10µl of sample or controls. Thermal cycling was performed with the QuantStudio™ 6 Pro on fast mode under the following conditions: uracil-N-glycosylase incubation at 25°C for 2 minutes, reverse transcription at 50°C for 15□minutes, an initial denaturation at 95°C for 2□minutes, followed by 45 cycles of 95°C for 3 seconds and 55°C for 30 seconds. All samples were analyzed in duplicates. Viral RNA copies were calculated from a standard curve constructed with the N gene positive control plasmid (Integrated DNA Technologies, USA).

### Statistical Analyses

Data analyses were completed using STATA version 13.0 (StataCorp, College Station, Texas, USA). Fisher’s exact test was used to compare categorical variables, while Mann-Whitney U test was used to compare continuous variables between patients with and without detectable virus to identify variables associated with viral shedding in respiratory aerosols. Kruskal-Wallis test was used to compare median viral loads of different respiratory activities within the subgroup of patients with detectable virus in respiratory aerosols. All statistical tests were two-sided and a p-value of <0.05 was considered significant.

## Results

Twenty-three patients were enrolled in the study, including 1 patient who withdrew before sample collection. Among the 22 participants, 19 (86%) were male, with median age of 38 years (range 23– 66). Five (23%) were asymptomatic (never developed symptoms). Thirteen (59%) emitted detectable levels of SARS-CoV-2 RNA in respiratory aerosols (Table 1), including 3 asymptomatic patients and 1 presymptomatic patient. SARS-CoV-2 copies emitted per expiratory activity per participant (30-minute breathing, 15-minute talking, or 15-minute singing) ranged from 63–5,821 viral N gene copies. Age, sex, virus variant type, clinical symptoms, presence of SARS-CoV-2 antibody at diagnosis, and Ct value of clinical sample at diagnosis, were not significantly different between patients with and without detectable viral RNA in respiratory aerosols (Table 2). However, the median day of illness was significantly different: patients with detectable viral RNA in aerosols were earlier in the course of illness (median day of illness of 3 versus 5, p-value=0.025).

**Table 1.** Severe acute respiratory syndrome coronavirus 2 (SARS-CoV-2) in respiratory aerosols emitted by coronavirus disease 2019 (COVID-19) patients in Singapore, February – April 2021

| Participant | Symptoms | Day of illness<sup>a</sup> | Clinical Ct value<sup>b</sup> | SARS-CoV-2 serology | Aerosolized SARS-CoV-2 RNA copies emitted<sup>c</sup> |  |  |  | SARS-CoV-2 variant |
| --- | --- | --- | --- | --- | --- | --- | --- | --- | --- |
|  |  |  |  |  | Breathing<sup>d</sup> | Talking<sup>e</sup> | Singing<sup>f</sup> | Total |  |
| 1 | Sore throat, rhinorrhea, anosmia, fever | 6 | 14.3 | Positive | ND | ND | ND | -- | Failed WGS |
| 2 | Rhinorrhea, anosmia | 7 | 16.6 | Negative | ND | ND | ND | -- | Alpha (B.1.1.7) |
| 3 | Sore throat, chronic cough | 9 | 30 | Negative | ND | ND | ND | -- | Non-VOC/VOI |
| 4 | Rhinorrhea, anosmia, cough, SOB | 2 | 19.4 | Negative | ND | 417 | ND | **417** | Alpha (B.1.1.7) |
| 5 | Asymptomatic | 5 (day of diagnosis) | 22.4 | Negative | ND | 234.5 | 135.2 | **369.7** | Non-VOC/VOI |
| 6 | Sore throat, rhinorrhea | 1 | 13.2 | Negative | ND | 79.9 | 713.6 | **793.5** | Beta (B.1.351) |
| 7 | Asymptomatic | 3 (day of diagnosis) | 32.9 | Positive | ND | ND | ND | -- | Failed WGS |
| 8 | Slight sore throat and rhinorrhea (due to swab test), fever | 5 | 16.5 | Negative | ND | ND | ND | -- | Non-VOC/VOI |
| 9 | Rhinorrhea, cough | 3 (day of diagnosis; 2 days pre-symptom onset) | 15.4 | Positive | ND | 908.2 | ND | **908.2** | Beta (B.1.351) |
| 10 | Sore throat | 4 | 16.1 | Negative | 63.5 | 310.9 | 1811.7 | **2186.1** | Non-VOC/VOI |
| 11 | Rhinorrhea, fever | 8 | 17 | Negative | ND | ND | 154.4 | **154.4** | Beta (B.1.351) |
| 12 | Fever, dry throat | 3 | 15.4 | Negative | 227.6 | 4336 | 4277.9 | **8841.5** | Alpha (B.1.1.7) |
| 13 | Fever | 2 | 19.2 | Negative | 140.9 | 733 | ND | **874** | Beta (B.1.351) |
| 14 | Fever, dry cough | 4 | 15.1 | Negative | ND | ND | ND | -- | Alpha (B.1.1.7) |
| 15 | Fever | 5 | 16.8 | Negative | 442.1 | 1356.5 | 978.8 | **2777.5** | Kappa (B.1.617.1) |
| 16 | Fever | 3 | 14.7 | Negative | 224.2 | 1373.3 | 5821.4 | **7419** | Beta (B.1.351) |
| 17 | Asymptomatic | 2 (day of diagnosis) | 14.5 | Positive | ND | ND | 143.6 | **143.6** | Kappa (B.1.617.1) |
| 18 | Asymptomatic | 3 (day of diagnosis) | 15.3 | Negative | 550.3 | 477.9 | 1216.1 | **2244.3** | Kappa (B.1.617.1) |
| 19 | Asymptomatic | 3 (day of diagnosis) | 14.4 | Negative | ND | ND | ND | -- | Beta (B.1.351) |
| 20 | Diarrhea, intermittent blocked nose | 5 | 19.5 | Positive | ND | ND | ND | -- | Beta (B.1.351) |
| 21 | Sore throat, fever, body ache | 5 | 16 | Negative | 310.5 | 2428.7 | 1162.3 | **3901.4** | Delta (B.1.617.2) |
| 22 | Rhinorrhea, fever, cough | 9 | 17.7 | Negative | ND | ND | ND | -- | Beta (B.1.351) |

ND = none detected; SOB = shortness of breath; WGS = whole genome sequencing; VOC = variant of concern; VOI = variant of interest
<sup>a</sup>On aerosol sample collection day; for symptomatic patients, day one of illness was defined as the day symptoms began; for asymptomatic and presymptomatic patients, day one of illness was defined as the day of diagnosis (day of the first PCR-positive clinical sample)
<sup>b</sup>PCR cycle threshold value from patient's diagnostic sample
<sup>c</sup>Viral N gene copies per expiratory activity
<sup>d</sup>30 minutes of tidal breathing
<sup>e</sup>15 minutes of talking with brief pauses
<sup>f</sup>15 minutes of continuous singing

**Table 2.**
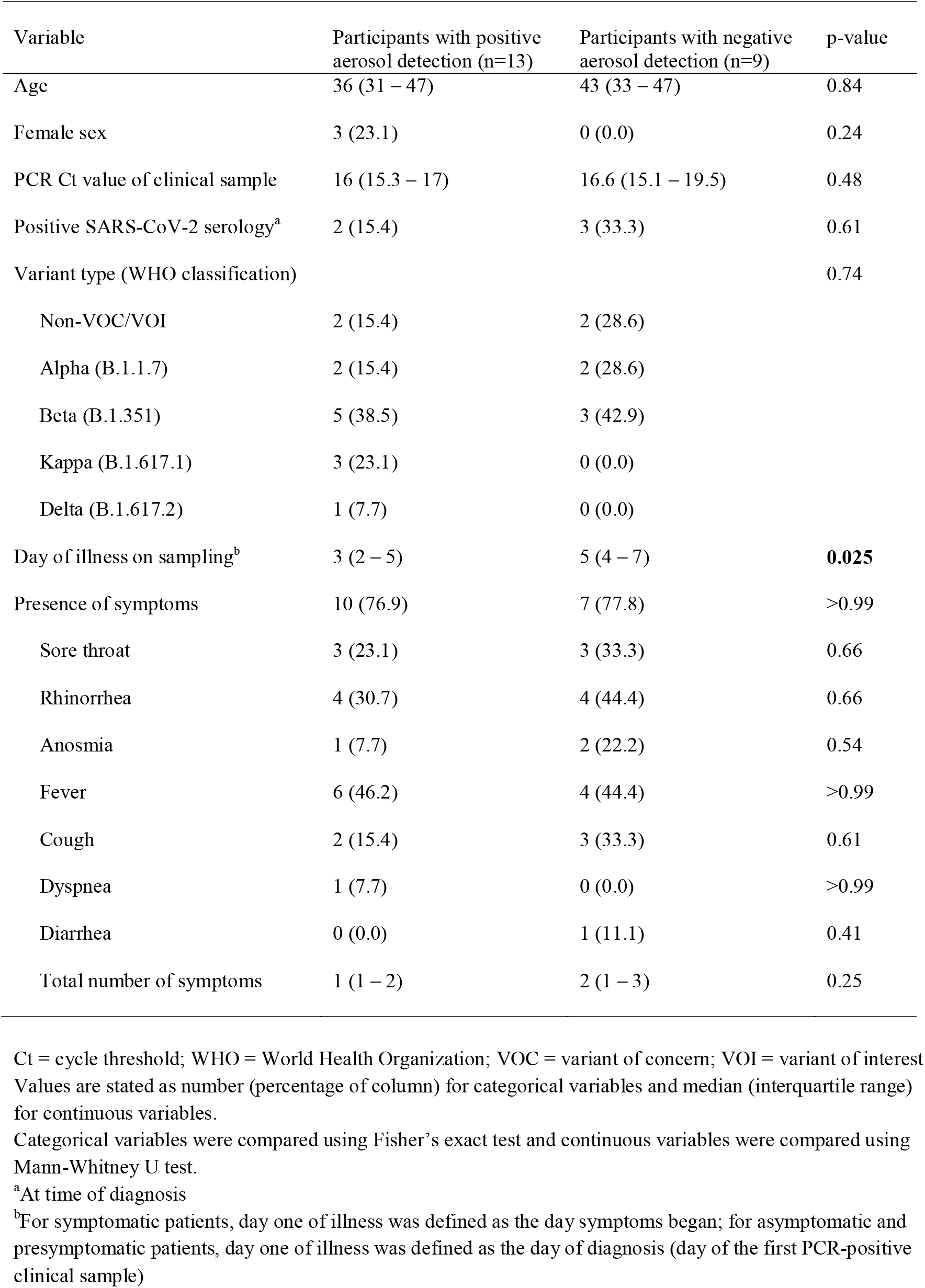
Comparison of variables between COVID-19 patients with and without detectable virus in respiratory aerosols

Six participants (27%) emitted detectable levels of SARS-CoV-2 RNA from all the expiratory activities. Two (9%) emitted detectable levels only from fine speech aerosols. Another two emitted detectable levels only from singing. No patients were observed to have sneezed during sample collection; however, two participants were observed to be coughing. Participant 4, who emitted 417 RNA copies in fine speech aerosols, was coughing during talking and singing. Participant 22 coughed frequently during all three activities but did not emit detectable viral RNA. Altogether, most SARS-CoV-2 RNA copies were emitted by singing (53%), followed by talking (41%) and breathing (6%) (Table 3).

**Table 3.** Sum total of viral RNA loads emitted in coarse and fine respiratory aerosols, for a sub-group of COVID-19 patients with detectable SARS-CoV-2 in respiratory aerosols (n=13)

|  | Coarse fraction | Fine fraction | Total (% of column) |
| --- | --- | --- | --- |
| Three expiratory activities | 4527.3 (14.6) | 26,503 (85.4) | **31,030.3** |
| Breathing<sup>a</sup> | 897 (45.8; 2.9) | 1,062.3 (54.2; 3.4) | 1959.3 (6.3) |
| Talking<sup>b</sup> | 868.4 (6.9; 2.7) | 11,787.5 (93.1; 38) | 12,655.9 (40.8) |
| Singing<sup>c</sup> | 2,762 (16.8; 9) | 13,653.2 (83.2; 44) | 16,415.5 (52.9) |

All values expressed as: viral N gene copies (percentage of row; percentage of **overall total**), unless otherwise noted.
<sup>a</sup>30 minutes of tidal breathing
<sup>b</sup>15 minutes of talking with brief pauses
<sup>c</sup>15 minutes of continuous singing

Viral loads in respiratory aerosols differed significantly between the three activities. Comparing only patients with detectable SARS-CoV-2 RNA in aerosols (n=13), the median number of viral N gene copies generated during singing was 713.6 (IQR 135.1–1216.1), compared to 477.9 (IQR 234.5–1356.6) for talking, and 63.5 (0–227.6) for breathing (Kruskal-Wallis test, p=0.026). Further comparison revealed that this difference remained significant for fine aerosols, but not for coarse aerosols (Table 4). Altogether, fine aerosols (≤5µm in diameter) constituted 85.4% of the total viral RNA load detected in our study.

**Table 4.**
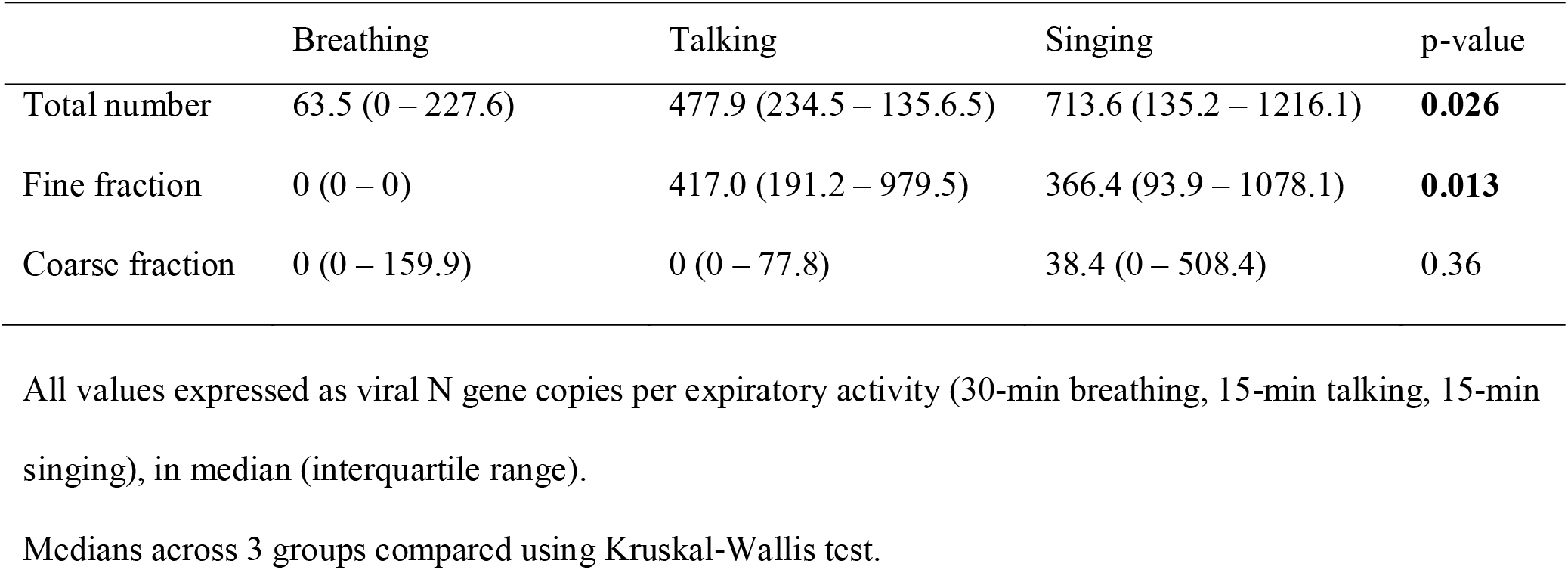
Median viral RNA loads emitted for each expiratory activity, in a sub-group of COVID-19 patients with detectable SARS-CoV-2 in respiratory aerosols (n=13)

### SARS-CoV-2 Variants

Sixteen participants (73%) were infected with a SARS-CoV-2 variant of concern (VOC) or variant of interest (VOI) during our study (Table 1). Due to the small number of non-VOC/VOI variants, aerosol shedding patterns related to SARS-CoV-2 variant type could not be determined.

### SARS-CoV-2 Culture

Virus cultures were negative after two consecutive passages.

## Discussion

Our study demonstrates that SARS-CoV-2 can be aerosolized in the absence of coughing, sneezing, and aerosol-generating medical procedures. More than half of our study participants emitted detectable levels of SARS-CoV-2 RNA in respiratory aerosols, including 3 asymptomatic patients and 1 presymptomatic patient. Patients earlier in illness were more likely to emit detectable levels of virus, which is congruent with studies demonstrating higher viral loads in clinical samples in early illness [17]. Although the overall viral RNA loads were relatively low, they differed significantly between breathing, talking, and singing, with singing generating the most virus in aerosols, and breathing generating the least. Overall, 85% of the total viral load was emitted in fine aerosols (≤5µm in diameter) when compared to coarse aerosols (>5µm in diameter), which is consistent with the observation that smaller particles (0.65–4.7µm) account for 77–79% of total virus particles shed by experimentally infected cynomolgus macaques [15]. Our results demonstrate the potential for fine respiratory aerosols to play an important role in community transmission of SARS-CoV-2, which is in agreement with other expert views suggesting that SARS-CoV-2 transmission events are driven by the airborne route [18], and could explain the difficulty in containing the virus. Our results support the calls for proper respiratory protection, airflow patterns, ventilation, filtration, and safe airborne disinfection, particularly in indoor environments [19], to reduce exposure to SARS-CoV-2 in fine aerosols – albeit live virus could not be isolated.

While it has been previously shown that COVID-19 patients can emit infectious virus-laden aerosols into their environments [5, 20], most environmental SARS-CoV-2 sampling studies have been unable to mechanically retrieve and isolate viable virus from ambient air in the vicinity of COVID-19 patients [21]. Hence, the infectious proportion of virus emitted from patient expiration remains unclear. In our study, the inability to isolate viable virus from respiratory aerosol samples collected directly from patients (not from their environments) is likely related to the low viral load in our samples compared to those generally found in culturable clinical samples. Our study was limited in that respiratory swabs were not collected on the day of aerosol sampling for comparison of culturability. However, studies have reported that for clinical SARS-CoV-2 samples, viral loads of 10^5^ to 10^6^ genome copies/mL are required for isolation of SARS-CoV-2 *in vitro* [22]. Our sampling methodology yielded viral RNA loads below 10^3.8^ genome copies per sample, suggesting that increased sampling duration is needed to reach culturable virus levels. However, critical mutations in certain SARS-CoV-2 variants can augment virus infectivity [23], e.g., some patients infected with the Delta variant demonstrate higher viral loads in their respiratory swabs [24]. These SARS-CoV-2 variants, especially Delta [24], can cause a higher secondary attack rate than older strains [25] and may be more successfully cultured from aerosol samples in future studies, especially if patients are sampled during the short window of enhanced viral shedding [26, 27]. More studies are warranted to test this hypothesis given that only 4 study participants were infected with non-VOC/VOI variants, and only one with Delta. Thus, aerosol shedding patterns between early and new SARS-CoV-2 strains could not be compared. Additionally, for virus culture in our study, we did not employ Vero E6 cells expressing the transmembrane serine protease 2 (TMPRSS2) which can bind and cleave SARS-CoV-2 spike protein more efficiently and facilitate early surface-mediated cell entry and viral fusion [28, 29]. Although SARS-CoV-2 from saliva and respiratory swabs can be isolated using classical Vero E6 cells, a more sensitive culture assay using Vero E6 TMPRSS2 cells may be superior for culturing virus from patient aerosol samples. Human bronchial epithelial cells may also be more susceptible to infection with wildtype viruses than Vero cells [23]. Further efforts to identify optimal culture methods for exhaled breath and environmental samples are warranted.

We observed that patients earlier in illness were more likely to emit detectable levels of virus in aerosols, which is in line with a recent non-human primate model indicating that SARS-CoV-2 aerosol shedding is substantially reduced 4 days post-infection when compared to 2 days post-infection [15], and concurs with the higher viral loads and greater infectivity observed in human clinical samples collected early in illness [17]. Additionally, neutralizing antibodies start to appear in COVID-19 patients five days post-symptom onset [30], which may reduce and neutralize virus that is shed, preventing isolation in cell culture. Although 17 participants (77%) were seronegative at diagnosis (Table 1), a serology test nearer the sampling day would have been a better indicator of infectiousness during aerosol sampling. Although 12 (55%) were sampled with the G-II machine within 5 days post-symptom onset (plus Participant 9, sampled 2 days pre-symptom onset), we failed to isolate viable virus, suggesting that participants might need to be sampled at an earlier stage of infection, or for longer durations. Recent data suggest that only 2% of infected individuals carry 90% of the total viral load circulating in a population at any given time [26]. This implies that only 1 in 50 active cases at any given time would be expected to have high viral loads in exhaled breath. The likelihood of capturing such cases was limited by our small sample size. Thus, researchers must work with contact tracers to proactively isolate and strategically sample large numbers of close contacts of individuals recently infected with SARS-CoV-2 to capture the most accurate data on viral shedding in the community, for which research gaps remain.

Viral RNA loads differed significantly between each of the three activities performed. The most virus copies were emitted by singing, followed by talking and breathing. To our knowledge, this is the first study to quantify SARS-CoV-2 in aerosols generated by singing. Our results support existing laboratory simulation data [31, 32], and can explain the many airborne SARS-CoV-2 outbreaks involving singing [8, 9, 33-35]. The lower number of virus copies detected in speech aerosols may be partially attributed to our study design. While participants sang continuously for 15 minutes, the talking portion of our study was designed to simulate a 15-minute conversation at rest (with pauses) rather than a continuous 15-minute speech/lecture. Higher concentrations of aerosols are also generated by singing compared to talking, with loudness having a large effect on the number of aerosols produced [31, 32, 36]. Individuals who generate an above-average amount of aerosols (known as “super-emitters”) also exist, but it is unclear what causes this phenomenon [37].

Interestingly, a small number of individuals produce more aerosols from breathing when compared to talking [32], which may partially explain the asymptomatic participant in our study who emitted more SARS-CoV-2 from breathing rather than talking. The physiological or experimental reasons underlying this observation are unclear.

Our results underscore the importance of reducing exposure to fine respiratory aerosols through non-pharmaceutical interventions (NPIs), such as universal masking, physical distancing, and increased room ventilation during the COVID-19 pandemic. Additionally, portable high efficiency particulate air (HEPA) cleaners in indoor environments can reduce exposure to exhaled respiratory aerosols by up to 90% in combination with universal masking, and up to 65% without universal masking [38], indicating that a multilayered approach of control measures is most effective at decreasing the risk of airborne SARS-CoV-2 transmission. Other NPIs include upper-room ultraviolet air disinfection, and the use of fans to control airflow patterns within a space. In singing situations, safe distancing among singers and averting and filtering airflow from choir to audience (e.g., by deploying air curtains), are important considerations. For situations involving talking, determining airflow patterns and minimizing exposure through seating and furniture configurations, distancing, and air movement alteration (such as fans, including desk fans) would be practical options [39, 40].

## Conclusion

Fine aerosols (≤5μm) produced by talking and singing contain more SARS-CoV-2 than coarse aerosols (>5μm) and may play a significant role in the transmission of SARS-CoV-2. Thus, exposure to fine aerosols should be mitigated, especially in indoor environments where airborne transmission of SARS-CoV-2 is most likely to occur. While COVID-19 patients shed detectable levels of SARS-CoV-2 RNA in respiratory aerosols, culturing SARS-CoV-2 from patient aerosol samples remains challenging. Careful focus is needed on sampling methodology and duration, infectiousness of patients during sampling, and virus culture methodology. Whether isolating viable virus in respiratory aerosols can be more easily accomplished from sampling patients infected with emerging SARS-CoV-2 variants is an important enquiry for future investigations. Minimizing airborne transmission by altering or averting direct airflow exposure in singing and speech situations within indoor environments may be important practical options to adopt.

## Data Availability

Data are available from coauthors upon reasonable request.

## Data Availability

Data can be provided upon reasonable request.

## Acknowledgment

We thank Tan Chorh Chuan and Leo Yee-Sin for their administrative support, and Julian Tang for his virology insights. We also thank Chandra Sekhar and David Kok Wai Cheong for assembling the G-II machine, and Somayeh Youssefi, Jacob Bueno De Mesquita and Jovan Pantelic for guiding the G-II assembly and operation. We thank Raymond Lin and Lin Cui for sharing whole genome sequencing data. We also thank Margaret Soon, Phoon Long Yoke, Loh Kyun Yen, Pang Jia Xin, and all nursing, infection control, and operational staff at the National Centre for Infectious Diseases for their support. Lastly, we thank the NUS BSL-3 Core Facility team for their support in BSL-3 procedures. We gratefully acknowledge the University of Maryland for the availability of the G-II machine for the study.

## Ethics Statement

This study was approved by the National Healthcare Group Domain Specific Review Board, reference number 2020/01113. Written informed consent was obtained from all study participants.

## References

1. Pan Y, Zhang D, Yang P, Poon LL, Wang Q. Viral load of SARS-CoV-2 in clinical samples. The Lancet infectious diseases 2020; 20(4): 411–2.

2. Zou L, Ruan F, Huang M, et al. SARS-CoV-2 viral load in upper respiratory specimens of infected patients. New England Journal of Medicine 2020; 382(12): 1177–9.

3. Chia PY, Coleman KK, Tan YK, et al. Detection of air and surface contamination by SARS-CoV-2 in hospital rooms of infected patients. Nature communications 2020; 11(1): 1–7.

4. Liu Y, Ning Z, Chen Y, et al. Aerodynamic analysis of SARS-CoV-2 in two Wuhan hospitals. Nature 2020; 582(7813): 557–60.

5. Lednicky JA, Lauzard M, Fan ZH, et al. Viable SARS-CoV-2 in the air of a hospital room with COVID-19 patients. International Journal of Infectious Diseases 2020; 100: 476–82.

6. Stadnytskyi V, Anfinrud P, Bax A. Breathing, speaking, coughing or sneezing: What drives transmission of SARSLCoVL2? Journal of Internal Medicine 2021.

7. Johansson MA, Quandelacy TM, Kada S, et al. SARS-CoV-2 transmission from people without COVID-19 symptoms. JAMA network open 2021; 4(1): e2035057–e.

8. Hamner L. High SARS-CoV-2 attack rate following exposure at a choir practice—Skagit County, Washington, March 2020. MMWR Morbidity and mortality weekly report 2020; 69.

9. Miller SL, Nazaroff WW, Jimenez JL, et al. Transmission of SARSLCoVL2 by inhalation of respiratory aerosol in the Skagit Valley Chorale superspreading event. Indoor air 2021; 31(2): 314–23.

10. Zhang N, Chen X, Jia W, et al. Evidence for lack of transmission by close contact and surface touch in a restaurant outbreak of COVID-19. Journal of Infection 2021.

11. Milton DK. A Rosetta Stone for understanding infectious drops and aerosols. Oxford University Press US, 2020.

12. Milton DM. What was the primary mode of smallpox transmission? Implications for biodefense. Frontiers in cellular and infection microbiology 2012; 2: 150.

13. Bixler SL, Stefan CP, Jay A, et al. Aerosol Exposure of Cynomolgus Macaques to SARS-CoV-2 Results in More Severe Pathology than Existing Models. bioRxiv 2021.

14. Woolsey C, Borisevich V, Prasad AN, et al. Establishment of an African green monkey model for COVID-19 and protection against re-infection. Nature immunology 2021; 22(1): 86–98.

15. Zhang C, Guo Z, Zhao Z, et al. SARS-CoV-2 Aerosol Exhaled by Experimentally Infected Cynomolgus Monkeys. Emerging Infectious Diseases 2021; 27(7): 1979–81.

16. McDevitt JJ, Koutrakis P, Ferguson ST, et al. Development and performance evaluation of an exhaled-breath bioaerosol collector for influenza virus. Aerosol Science and Technology 2013; 47(4): 444–51.

17. Kim M-C, Cui C, Shin K-R, et al. Duration of culturable SARS-CoV-2 in hospitalized patients with Covid-19. New England Journal of Medicine 2021; 384(7): 671–3.

18. Greenhalgh T, Jimenez JL, Prather KA, Tufekci Z, Fisman D, Schooley R. Ten scientific reasons in support of airborne transmission of SARS-CoV-2. The lancet 2021; 397(10285): 1603–5.

19. Qian H, Miao T, Liu L, Zheng X, Luo D, Li Y. Indoor transmission of SARSLCoVL2. Indoor Air 2021; 31(3): 639–45.

20. Santarpia JL, Rivera DN, Herrera VL, et al. Aerosol and surface contamination of SARS-CoV-2 observed in quarantine and isolation care. Scientific reports 2020; 10(1): 1–8.

21. Ong SWX, Tan YK, Coleman KK, et al. Lack of viable severe acute respiratory coronavirus virus 2 (SARS-CoV-2) among PCR-positive air samples from hospital rooms and community isolation facilities. Infection Control & Hospital Epidemiology 2021: 1–6.

22. Huang C-G, Lee K-M, Hsiao M-J, et al. Culture-based virus isolation to evaluate potential infectivity of clinical specimens tested for COVID-19. J Clin Microbiol 2020; 58(8): e01068–20.

23. Pohl MO, Busnadiego I, Kufner V, et al. SARS-CoV-2 variants reveal features critical for replication in primary human cells. PLoS biology 2021; 19(3): e3001006.

24. Ong SWX, Chiew CJ, Ang LW, et al. Clinical and Virological Features of SARS-CoV-2 Variants of Concern: A Retrospective Cohort Study Comparing B. 1.1. 7 (Alpha), B. 1.315 (Beta), and B. 1.617. 2 (Delta). Available at SSRN: https://doiorg/102139/ssrn3861566.

25. Public Health England. SARS-CoV-2 variants of concern and variants under investigation in England. technical briefing 12 2021.

26. Yang Q, Saldi TK, Gonzales PK, et al. Just 2% of SARS-CoV-2-positive individuals carry 90% of the virus circulating in communities. Proceedings of the National Academy of Sciences 2021; 118(21).

27. Kim KS, Ejima K, Iwanami S, et al. A quantitative model used to compare within-host SARS-CoV-2, MERS-CoV, and SARS-CoV dynamics provides insights into the pathogenesis and treatment of SARS-CoV-2. PLoS biology 2021; 19(3): e3001128.

28. Hoffmann M, Kleine-Weber H, Schroeder S, et al. SARS-CoV-2 cell entry depends on ACE2 and TMPRSS2 and is blocked by a clinically proven protease inhibitor. cell 2020; 181(2): 271-80. e8.

29. Murgolo N, Therien AG, Howell B, et al. SARS-CoV-2 tropism, entry, replication, and propagation: Considerations for drug discovery and development. PLoS Pathogens 2021; 17(2): e1009225.

30. Long Q-X, Liu B-Z, Deng H-J, et al. Antibody responses to SARS-CoV-2 in patients with COVID-19. Nature medicine 2020; 26(6): 845–8.

31. Alsved M, Matamis A, Bohlin R, et al. Exhaled respiratory particles during singing and talking. Aerosol Science and Technology 2020; 54(11): 1245–8.

32. Gregson FK, Watson NA, Orton CM, et al. Comparing aerosol concentrations and particle size distributions generated by singing, speaking and breathing. Aerosol Science and Technology 2021; 55(6): 681–91.

33. Katelaris AL, Wells J, Clark P, et al. Epidemiologic evidence for airborne transmission of SARS-CoV-2 during church singing, Australia, 2020. Emerging infectious diseases 2021; 27(6): 1677.

34. Charlotte N. High rate of SARS-CoV-2 transmission due to choir practice in France at the beginning of the COVID-19 pandemic. Journal of Voice 2020.

35. Gu Y, Lu J, Su W, Liu Y, Xie C, Yuan J. Transmission of SARS-CoV-2 in the Karaoke Room: An Outbreak of COVID-19 in Guangzhou, China, 2020. Journal of Epidemiology and Global Health 2021; 11(1): 6.

36. Philip KE, Lewis A, Buttery SC, et al. Aerosol transmission of SARS-CoV-2: inhalation as well as exhalation matters for COVID-19. American journal of respiratory and critical care medicine 2021; 203(8): 1041–2.

37. Asadi S, Wexler AS, Cappa CD, Barreda S, Bouvier NM, Ristenpart WD. Aerosol emission and superemission during human speech increase with voice loudness. Scientific reports 2019; 9(1): 1–10.

38. Lindsley WG. Efficacy of Portable Air Cleaners and Masking for Reducing Indoor Exposure to Simulated Exhaled SARS-CoV-2 Aerosols—United States, 2021. MMWR Morbidity and Mortality Weekly Report 2021; 70.

39. Li W, Chong A, Hasama T, et al. Effects of ceiling fans on airborne transmission in an air-conditioned space. Building and Environment 2021; 198: 107887.

40. Pantelic J, Sze-To GN, Tham KW, Chao CY, Khoo YCM. Personalized ventilation as a control measure for airborne transmissible disease spread. Journal of the Royal Society Interface 2009; 6(Suppl_6): S715–S26.

